# From frail to fit after allogeneic hematopoietic cell transplantation: Scope of the problem and a strength-based solution

**DOI:** 10.1101/19010397

**Authors:** Jason Sweetnam, Eric Twohey, Sasha Skendzel, John Stingle, Mukta Arora, Austin Baraki, Qing Cao, Sonya Grillo, Shernan G. Holtan

## Abstract

**Background:** Frailty is a common but underdiagnosed syndrome among long-term survivors of allogeneic hematopoietic cell transplantation (HCT). Conditions such as malnutrition, fatigue, and weakness may suggest frailty despite patients not receiving a formal diagnosis. Furthermore, the optimal interventions to overcome frailty in long-term survivors of allogeneic HCT is not yet established.

**Patients and Methods:** This study consists of a retrospective and prospective component. First, we completed a retrospective review using diagnosis data from the electronic medical record to estimate the prevalence of components of frailty in 1077 recipients of allogeneic HCT through 5 years post-transplant. Second, we developed a community-based pilot study of strength training for long-term survivors of allogeneic HCT that addressed several common barriers exercise engagement and adherence. Four allogeneic HCT recipients (2 males, 2 females) and 4 controls (2 males, 2 females) completed the strength training pilot study, consisting of a baseline assessment, 10 weeks of personalized and supervised strength programming at least once weekly as a group, and an end-of-study assessment to measure progress in strength, body composition, and a standardized measure of self-efficacy.

**Results:** Despite a lack of a formal diagnosis, approximately 80% of HCT recipients in this series received a diagnosis of a component of frailty (weakness, fatigue, or malnutrition), and over 1/3 of HCT recipients had symptoms extending beyond 1 year. Over the course of the strength training pilot study, both pre-frail/frail allogeneic HCT recipients and healthy controls doubled their total strength, lost body fat, and gained muscle mass. Self-efficacy improved from baseline in allogeneic HCT recipients after the strength training pilot program.

**Discussion:** Based upon the prevalence of frailty-related diagnoses among long-term survivors of HCT, a comprehensive mitigation strategy needs to be developed for this population. A community-based strength training program that includes a personalized component, a group setting, and caregiver/partner involvement appears feasible and overcomes several established barriers to exercise engagement and adherence.

## Introduction

Frailty is a clinical syndrome of diminished physiologic reserve and low resistance to stressors affecting approximately 10% of the hematopoietic cell transplant (HCT) population.(*1*) Frailty can manifest as weakness, low physical activity, exercise intolerance, unintentional weight loss, self-reported exhaustion, and low-self efficacy.(*2, 3*) While its manifestations are common and undoubtedly recognized as expected or “normal” to some extent, frailty is likely underdiagnosed in HCT recipients. Conditions such as malnutrition, fatigue, and weakness may suggest frailty despite patients not receiving a formal diagnosis. Because aspects of frailty are so common after HCT, its root causes, including potential anabolic resistance (*4*), may not be adequately addressed by physicians specializing in HCT. Therefore, there is an unmet need to both increase the recognition of frailty by the medical team and to design and test interventions to mitigate it.

The optimal interventions to prevent or mitigate frailty in the allogeneic HCT population are not yet known. However, strength-based training programs directly address multiple domains of frailty and have shown success in other clinical settings, including community-dwelling elderly (*5*), those residing in long-term care facilities (*6*), and after lung transplantation (*7*). We have 2 goals outlined in this paper: (1) estimate the prevalence of components of frailty in 1077 recipients of allogeneic HCT through 5 years post-transplant using diagnosis data from the electronic medical record, and (2) perform a community-based pilot study of strength training in long-term survivors of allogeneic HCT. The results herein are useful to inform the development of a larger strength-focused longitudinal training program to reverse frailty and improve quality of life after allogeneic HCT.

## Patients and Methods

We performed this study in two parts. We first performed a retrospective review to estimate frailty from medical diagnosis codes abstracted from the electronic medical record. We subsequently performed a pilot study to improve frailty in cancer survivors. We describe details of each cohort below.

### Retrospective Cohort

In order to estimate the prevalence of frailty through 5 years post-HCT, we retrieved both International Classification of Diseases (ICD)9 and ICD10 codes for malnutrition, fatigue, and weakness from our electronic medical record system spanning 3 post-transplant periods: Day 0 – 100, Day 101 – 365, and 1 – 5 years. Patients (N=1077) in this cohort were adults age 18 years or greater undergoing HCT at the University of Minnesota between 2000-2016. We adjusted for mortality by taking into consideration the death rate in each time period. A patient could be counted for a diagnosis of malnutrition, fatigue, and/or weakness in the time period if they survived at all during that time period.

### Strength Training Pilot Study

We designed a pilot study of community-based strength training designed to address frailty. Participants (N=4) were required to be at least 18 months post-HCT, meet criteria for pre-frailty or frailty (all had low energy expenditure and self-reported exhaustion), and have a healthy exercise partner (spouse or other relative or friend) who could serve as a control (N=4). Participants in our pilot study received an assessment by a National Academy of Sports Medicine (NASM)-certified trainer prior to design of a personalized strength training program during week 1. During weeks 2-11, each participant completed their personalized program at least weekly as a group under the supervision of the trainer on Saturday mornings, and they were encouraged but not required to complete additional training sessions as individuals during the other days of the week. These additional training sessions were unsupervised but followed the same protocol as the supervised sessions. Each participants’ personalized strength training program consisted of three lower body exercises, four upper body exercises, three core exercises, and one aerobic test (example provided in Figure 1). Prescriptions for all exercises were determined by the number of repetitions at a given weight required to approach muscular failure in the initial assessment. The large majority of exercises were performed on Free Motion® machines. These machines have a rotating cable system so that individuals with different body types and proportions can use the same machines without being limited to a fixed motor pattern or range of motion.

**Figure 1.**
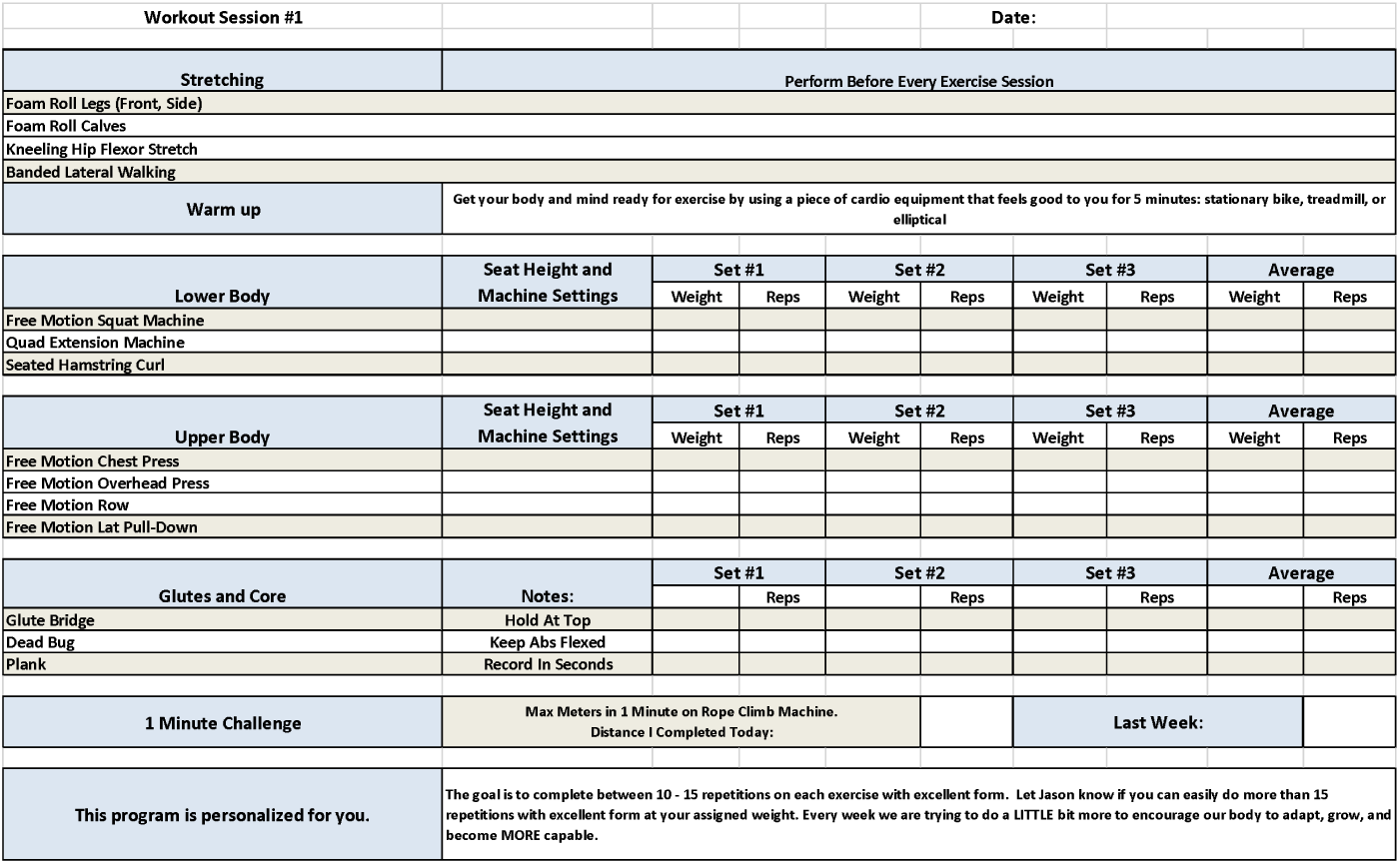
Example strength training prescription and training log.

During the course of the study, specific exercises and their loads were progressed based on the abilities of each participant. In assessing the progress of individuals during week 12 (final week of study), the estimate of improvement was based on the change in relative strength. This metric consists of changes in amount of weight lifted (absolute strength) and the progress in the amount of repetitions completed (work capacity). All of the participants’ workouts (both those with and without a trainer) were recorded and collected, and the final results were calculated by comparing subject’s initial assessments with their final workouts. To assess progress over the trial period, we compared relative strength in each of the muscle groups comparing the initial and final sessions using descriptive statistics (average, range) without statistical tests due to the small sample size.

### Body Composition

We recorded body composition, including weight, percentage of body fat, and weight of muscle mass, at the initial and final sessions using an InBody® bioelectrical impedance device. Participants were counseled that the goal of the study was not to lose weight but rather to gain muscle mass. No formal dietary intervention was given, but participants learned about healthy eating habits with a particular focus on eating for muscular growth from a registered dietician once per month in a group setting.

### Self-Efficacy Assessment

The primary endpoint of the strength training pilot study was self-efficacy as determined by a 6-question survey adapted from the Stanford Patient Education Resource Center (*8*). Participants gave a numeric response on a scale of 1 (not confident at all) to 10 (totally confident) on questions regarding confidence to deal with fatigue, physical discomfort, emotional distress, managing health problems, reducing dependence on the healthcare system, and taking steps to improve health beyond medications.

### Ethical Considerations

The University of Minnesota Institutional Review Board approved both the retrospective medical record review and the strength training pilot study. The strength training pilot study is registered on ClinicalTrials.gov as NCT03609203.

## Results

### Estimation of frailty from diagnosis codes

Demographics of the retrospective cohort are detailed in Table 1. The median age of this cohort was 52 years, with a slight male predominance (58.6% of patients). Table 2 lists the overall frequency of diagnoses of weakness, malnutrition, and fatigue over the period of 2000-2016, and Figure 2 shows how these diagnoses overlap in individual patients across early, intermediate, and late post-HCT recovery. The majority (79.3%) of adult allogeneic HCT recipients had a diagnosis of malnutrition (Table 2), weakness (Table 3), and/or fatigue (Table 4) at some time post-HCT. Only 179 patients (21.7%) had none of these diagnoses at any time point in their post-transplant recovery. Across the 3 time periods, fatigue was the most frequent diagnosis, with 37% diagnosed with fatigue prior to day 100, 28% diagnosed between days 101 and 365, and 33% diagnosed between years 1-5. The frequency of malnutrition diagnoses decreased over time, with 22% prior to day 100, 18% between days 101-365, and 16% between years 1-5. In contrast, the frequency of muscle weakness diagnoses increased over time, with 6% diagnosed prior to day 100, 9% between days 101 – 365, and 11% between years 1-5. Patients with all 3 diagnoses ranged from 1.8% in the first 100 days, to 3.9% between days 101 – 365, to 4.2% between years 1-5.

**Table 1.**
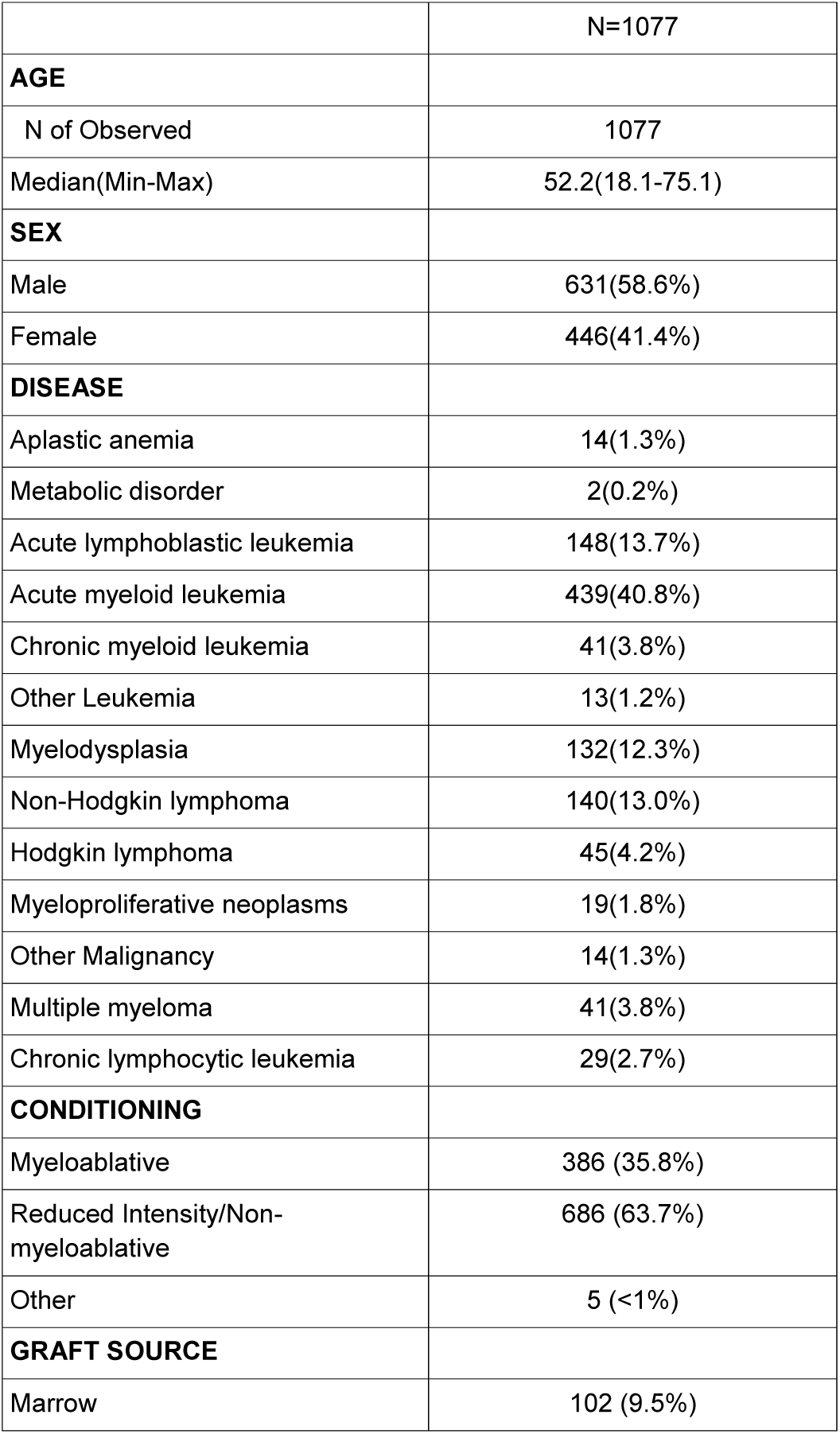
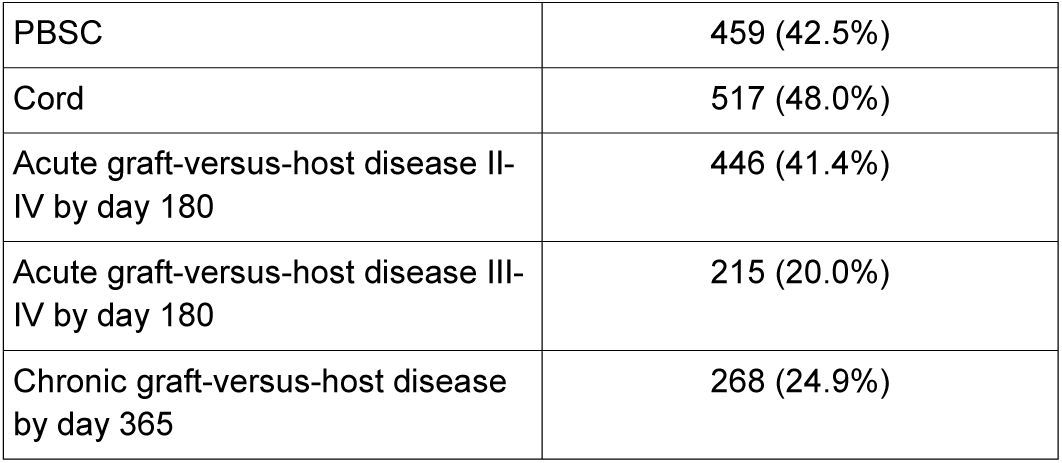
Retrospective cohort patient demographics.

|  |  |
| --- | --- |
|  | N=1077 |
| **AGE** |  |
| N of Observed | 1077 |
| Median(Min-Max) | 52.2(18.1-75.1) |
| **SEX** |  |
| Male | 631(58.6%) |
| Female | 446(41.4%) |
| **DISEASE** |  |
| Aplastic anemia | 14(1.3%) |
| Metabolic disorder | 2(0.2%) |
| Acute lymphoblastic leukemia | 148(13.7%) |
| Acute myeloid leukemia | 439(40.8%) |
| Chronic myeloid leukemia | 41(3.8%) |
| Other Leukemia | 13(1.2%) |
| Myelodysplasia | 132(12.3%) |
| Non-Hodgkin lymphoma | 140(13.0%) |
| Hodgkin lymphoma | 45(4.2%) |
| Myeloproliferative neoplasms | 19(1.8%) |
| Other Malignancy | 14(1.3%) |
| Multiple myeloma | 41(3.8%) |
| Chronic lymphocytic leukemia | 29(2.7%) |
| **CONDITIONING** |  |
| Myeloablative | 386 (35.8%) |
| Reduced Intensity/Non-myeloablative | 686 (63.7%) |
| Other | 5 (<1%) |
| **GRAFT SOURCE** |  |
| Marrow | 102 (9.5%) |
| PBSC | 459 (42.5%) |
| Cord | 517 (48.0%) |
| Acute graft-versus-host disease II-IV by day 180 | 446 (41.4%) |
| Acute graft-versus-host disease III-IV by day 180 | 215 (20.0%) |
| Chronic graft-versus-host disease by day 365 | 268 (24.9%) |

**Table 2.** Frequency of malnutrition diagnoses (raw data) abstracted from electronic medical record.

| Diagnosis Code | Code Type | Number of times diagnosed | Description |
| --- | --- | --- | --- |
| R63.0 | ICD10 | 523 | Anorexia |
| 783.0 | ICD9 | 443 | Anorexia |
| 783.21 | ICD9 | 236 | Loss of weight |
| R62.7 | ICD10 | 163 | Adult failure to thrive |
| R63.0 | ICD10 | 150 | Anorexia |
| 783.7 | ICD9 | 120 | Adult failure to thrive |
| 783.21 | ICD9 | 110 | Loss of weight |
| 783.7 | ICD9 | 98 | Adult failure to thrive |
| 783.0 | ICD9 | 92 | Anorexia |
| 783.3 | ICD9 | 91 | Feeding difficulties and mismanagement |
| R63.3 | ICD10 | 55 | Feeding difficulties |
| R62.7 | ICD10 | 55 | Adult failure to thrive |
| Z68.1 | ICD10 | 45 | Body mass index (BMI) 19 or less, adult |
| 783.7 | ICD9 | 30 | FAILURE TO THRIVE-ADULT |
| R64 | ICD10 | 29 | Cachexia |
| 799.4 | ICD9 | 28 | Cachexia |
| E44.0 | ICD10 | 25 | Moderate protein-calorie malnutrition |
| 263.0 | ICD9 | 16 | Malnutrition of moderate degree |
| R63.3 | ICD10 | 16 | Feeding difficulties |
| V85.0 | ICD9 | 16 | Body Mass Index less than 19, adult |
| E44.1 | ICD10 | 11 | Mild protein-calorie malnutrition |
| 783.3 | ICD9 | 9 | Feeding difficulties and mismanagement |
| E44.0 | ICD10 | 6 | Moderate protein-calorie malnutrition |
| 263.1 | ICD9 | 6 | Malnutrition of mild degree |
| 263.0 | ICD9 | 4 | Malnutrition of moderate degree |
| Z68.1 | ICD10 | 4 | Body mass index (BMI) 19 or less, adult |
| R62.51 | ICD10 | 3 | Failure to thrive (child) |
| E44.1 | ICD10 | 2 | Mild protein-calorie malnutrition |
| 783.41 | ICD9 | 2 | Failure to thrive |
| 783.3 | ICD9 | 1 | FEEDING PROBLEM |
| 799.4 | ICD9 | 1 | Cachexia |
| R64 | ICD10 | 1 | Cachexia |

**Table 3.** Frequency of weakness diagnoses (raw data) abstracted from electronic medical record.

| Diagnosis Code | Code Type | Number of times diagnosed | Description |
| --- | --- | --- | --- |
| 728.87 | ICD9 | 445 | Muscle weakness (generalized) |
| 728.87 | ICD9 | 377 | MUSCLE WEAKNESS-GENERAL |
| M62.81 | ICD10 | 223 | Muscle weakness (generalized) |
| M62.81 | ICD10 | 123 | Muscle weakness (generalized) |
| 728.87 | ICD9 | 118 | Muscle weakness (generalized) |
| M62.50 | ICD10 | 59 | Muscle wasting and atrophy, not elsewhere classified, unspecified site |
| M62.50 | ICD10 | 34 | Muscle wasting and atrophy, not elsewhere classified, unspecified site |
| M62.59 | ICD10 | 12 | Muscle wasting and atrophy, not elsewhere classified, multiple sites |
| M62.562 | ICD10 | 12 | Muscle wasting and atrophy, not elsewhere classified, left lower leg |
| M62.561 | ICD10 | 10 | Muscle wasting and atrophy, not elsewhere classified, right lower leg |
| M62.58 | ICD10 | 9 | Muscle wasting and atrophy, not elsewhere classified, other site |
| M62.552 | ICD10 | 5 | Muscle wasting and atrophy, not elsewhere classified, left thigh |
| M62.569 | ICD10 | 2 | Muscle wasting and atrophy, not elsewhere classified, unspecified lower leg |

**Table 4.** Frequency of fatigue diagnoses (raw data) abstracted from electronic medical record.

| Diagnosis Code | Code Type | Number of times diagnosed | Description |
| --- | --- | --- | --- |
| 780.79 | ICD9 | 2488 | Other malaise and fatigue |
| 780.79 | ICD9 | 1398 | Other malaise and fatigue |
| 780.79 | ICD9 | 817 | MALAISE AND FATIGUE NEC |
| R53.83 | ICD10 | 699 | Other fatigue |
| R53.83 | ICD10 | 340 | Other fatigue |
| R53.82 | ICD10 | 25 | Chronic fatigue, unspecified |
| 780.79 | ICD9 | 10 | MALAISE AND FATIGUE NEC |
| R53.82 | ICD10 | 6 | Chronic fatigue, unspecified |
| 780.54 | ICD9 | 5 | Hypersomnia, unspecified |
| 780.54 | ICD9 | 5 | Hypersomnia, unspecified |
| 780.53 | ICD9 | 4 | HYPERSOM W SLP APNEA NOS |
| 780.71 | ICD9 | 3 | Chronic fatigue syndrome |
| 780.71 | ICD9 | 3 | Chronic fatigue syndrome |
| G47.10 | ICD10 | 2 | Hypersomnia, unspecified |
| 780.54 | ICD9 | 2 | HYPERSOMNIA NOS |
| 780.79 | ICD9 | 1 | MALAISE AND FATIGUE NEC |
| G47.10 | ICD10 | 1 | Hypersomnia, unspecified |
| 780.54 | ICD9 | 1 | HYPERSOMNIA NOS |

**Figure 2.**
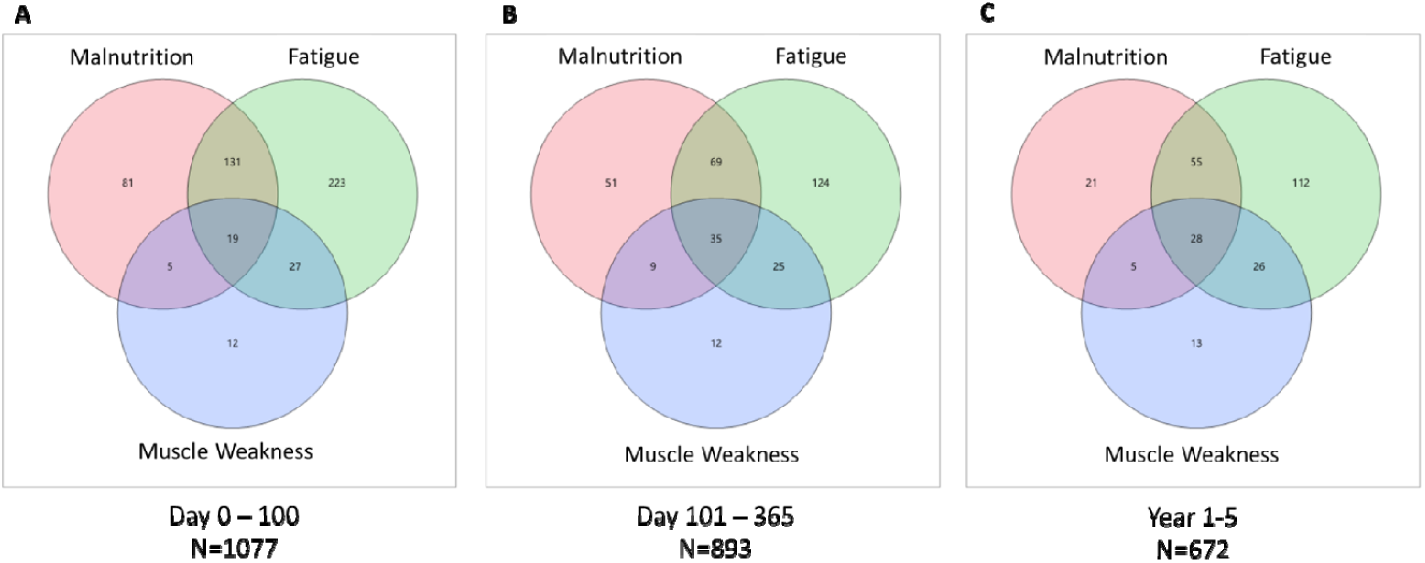
Number of patients with overlapping diagnoses of malnutrition, fatigue, and muscle weakness through 5 years post-allogeneic hematopoietic cell transplantation.

### Strength Training Pilot Study Outcomes

Four adult allogeneic HCT recipients (2 males, 2 females) and 4 controls (2 males, 2 females) completed the strength training pilot study, consisting of a baseline assessment, 10 weeks of personalized and supervised strength programming at least once weekly as a group, and an end-of-study assessment to measure progress. Participant engagement was excellent, there were no serious adverse events related to strength training. The average number of strength training sessions completed over 10 weeks by participant was 27 (range 17-34). Study participants doubled their strength on average after 10 weeks, as demonstrated by improvements of 194% in upper body exercises, and 199% in lower body exercises.

While this was not designed as a weight loss study, most participants lost fat mass. Average weight loss was 3.0 kilograms over 10 weeks, which reflected in an average change in body fat of 2.54% as assessed by bioelectrical impedance among study participants (Figure 3B). The average muscle gain as assessed by bioelectrical impedance in allogeneic HCT recipients was 1.4 kilograms throughout the study, with no net change in the control group (Figure 3C). However, one female outlier the healthy control group lost a total of 10.8 kg during the study.

**Figure 3.**
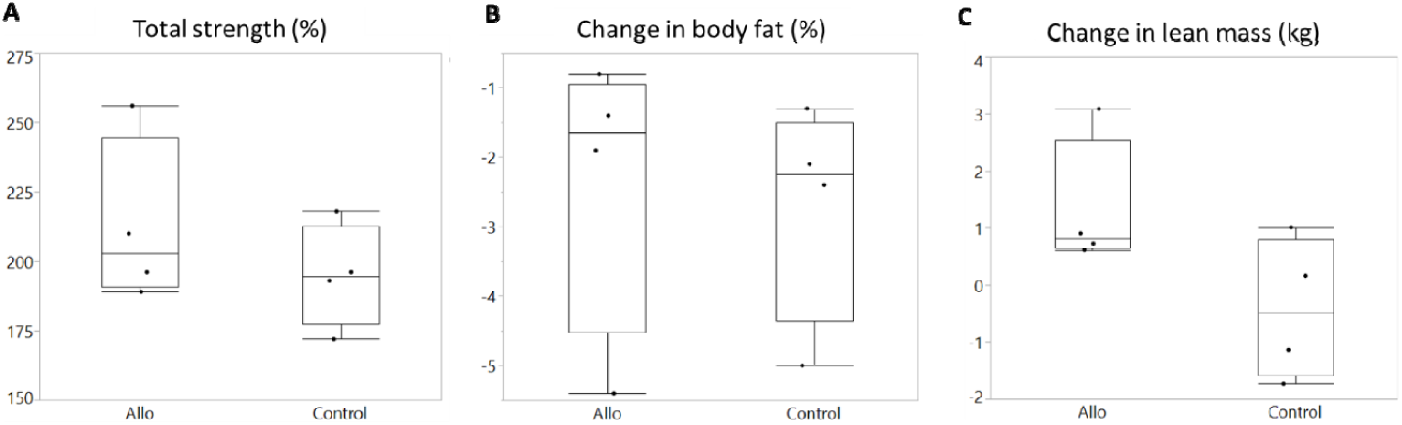
Comparison of percent change in (A) percent total strength compared to baseline, (B) percent body fat, (C) and lean body weight in kilograms between allogeneic hematopoietic cell transplant recipients (Allo, N=4) versus healthy controls (N=4).

We measured self-efficacy via 6-item survey at the start of the study and at its completion. Baseline self-efficacy was lower in recipients of allogeneic HCT than controls(Figure 4), but increased by an average of 10 points (mean 36.75 to 46.75) in allogeneic HCT recipients over the course of the study.

**Figure 4.**
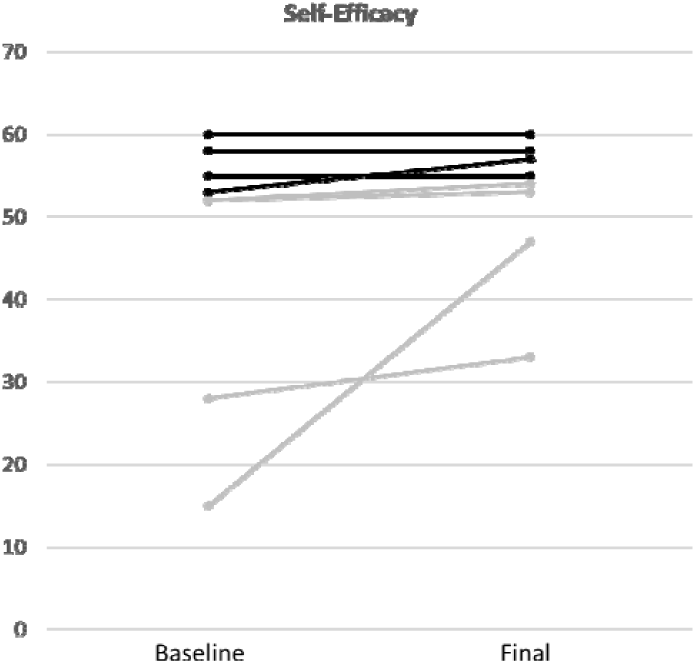
Self-efficacy at the start of the strength training intervention and at its completion (maximum score = 60 points) comparing recipients of allogeneic hematopoietic cell transplantation (gray) to controls (black).

## Discussion

Frailty is a commonly observed syndrome in the adult HCT population, although it is typically not diagnosed formally. Frailty does not have its own ICD-10 code; rather, the code R54 refers to “age-related physical debility” that includes frailty, but excludes sarcopenia and cognitive decline. Despite not receiving a formal diagnosis of frailty, approximately 80% of HCT recipients in this series received a diagnosis of a component of frailty (weakness, fatigue, or malnutrition), and over 1/3 of HCT recipients reported symptoms extending beyond 1 year. Sarcopenia affects 50% of HCT recipients even prior to HCT leading to an increased risk of late effects, poor quality of life, and early mortality.(*9*) Knowing this, and observing the frequency of frailty-related diagnoses in our series, a comprehensive mitigation strategy needs to be developed for long-term HCT survivors.

Recipients of HCT can carry several risk factors for frailty including advanced age, polypharmacy, lack of regular exercise, isolation, malnutrition, and unintentional weight loss (*1, 10*). Risks traditionally associated with life-threatening complications following SCT have been substantially mitigated by innovations in supportive care.(*11*) As a result, there has been a considerable increase in the number of SCT recipients living with the systemic effects of cancer and related treatment as a chronic condition.(*12*) In response, health organizations are now recommending survivorship interventions founded on the theoretical principles of the Chronic Care Model with a focus on programs targeting chronic-disease self-management.(*13*) Improving patient self-management behavior in the survivorship period has been the focus of multiple chronic disease programs.(*14-17*) Exercise has well-established beneficial effects including improved quality-of-life and a reduction in untoward cancer-related physical symptoms (*18*) and should thus be a key component of such self-management programs.

Perhaps the strongest predictor of health-related self-management behavior is self-efficacy.(*19*) As defined by Bandura, self-efficacy is the perceived ability or confidence to achieve a goal, which can be influenced by motivation and task mastery.(*20*) Increased self-efficacy among cancer survivors appears to have a relationship with exercise participation and adherence, with higher levels of self-efficacy associated with more exercise behaviors and vice versa.(*19, 21*) Understanding the critical role of this phenomenon in promoting and sustaining positive behavior change, we used the validated Chronic Disease Self-Management 6-Item scale to assess patient self-efficacy in this pilot study.(*8*) The scale measures several common domains of chronic disease including the management of symptoms and health related activities. Our pilot study found improvements in self-efficacy after the strength training intervention, but the overall level of self-efficacy still did not match that of healthy control participants. Self-efficacy may have continued to improve further with a longer study duration.

Our pilot study is one among many other larger studies that have investigated the role of exercise in recovery from HCT (Table 5). The results from previous interventions may be variable if the training stimulus was not personalized; either too small to induce muscular growth, or too large, putting individuals at risk of injury, excess fatigue, or pain. Factors that enhanced the feasibility of our pilot study is that the intervention was personalized and occurred in a commercial health club. While the participants were required to meet at least once weekly as a group at one of the club’s locations, the participants were able to exercise at a club closer to their home (if applicable) during the week. The required group participation also enhanced the feasibility and reduced expense, allowing the trainer to supervise 8 participants at once during the weekly sessions. The group participation also likely enhanced the sense of community among participants, as some remarked during the study that they felt less isolated than they had felt prior to starting the program. Finally, the partnering of healthy controls (caregiver, spouse, or other relative or friend) with the HCT recipients may have enhanced adherence to the program. We also wanted to involve caregivers in the intervention recognizing the impact of the HCT process on their lives.(*22*) We did not want to give them yet one more task to add to their list. Furthermore, involving the caregiver could impact the entire family dynamic toward adopting healthier lifestyle behaviors despite many perceived barriers when a loved one is going through the allogeneic HCT process.

**Table 5.** Prospective studies involving strength training in blood and marrow transplantation.

| **Study Design** | **N** | **Age** | **Intervention** | **Primary Endpoints** | **Results of Primary Endpoints** | **Secondary endpoints** | **Results of Secondary Endpoints** | **References** |
| --- | --- | --- | --- | --- | --- | --- | --- | --- |
| Randomized pilot study | 19 | Adult (ages 20-67) | Strength | Physical activity, muscle strength, fatigue, health status perceptions, quality of life | Significant decrease in fatigue with exercise group, no significant effects on all other primary outcomes | Not applicable | Not applicable | (23) |
| Non-randomized | 30 | Adolescents and adults | Strength | Muscle protein status and turnover- nitrogen balance, creatinine, 3-methylhistidine | Muscle protein sparing effect of exercise, significant decrease in creatinine excretion in controls only suggested muscle protein loss associated w/ inactivity | Arm muscle area | Correlated with energy but not protein intake | (24) |
| Randomized | 53 | Pediatric (ages 5-17) | Aerobic and Strength | 6-minute walk test (6MWT) | Intervention unfit group increased 6MWT, while unfit and fit control groups decreased. | Exercise frequency and intensity | Intervention fit group presented higher exercise frequency and intensity compared to intervention unfit | (25) |
| Randomized | 42 | Adult (ages 18-60) | Aerobic and Strength | VO2max | Significant increase | Muscle strength, functional performance, physical activity level, QOL, fatigue, psychological well-being and clinical outcomes | Significant effect on muscle strength: chest press, leg extension, right elbow flexor, right knee flexor, right knee extensor, and functional performance (stair test) | (26) |
| Randomized | 187 | Adults (ages 25-76) | Aerobic and strength in the context of Total Therapy for multiple myeloma | Fatigue, nighttime sleep, aerobic capacity | No statistical significance in nighttime sleep, 6MWT, or fatigue between groups | Not applicable | Not applicable | (27) |
| Randomized | 61 | Adults (ages 22-70) | Aerobic and Strength | Performance (50ft walk, 6MWT, functional reach, sit-to-stand, uniped stance) | No statistical significance found between supervised vs. unsupervised exercise groups | Brief Fatigue Inventory | No statistical significance | (28) |
| Randomized | 135 | Adults (ages 32-74) | Aerobic and Strength | # of transfusions during transplantation, # of attempts and total number of days of stem cell collection, time to recovery after transplant, response to intensive therapy | Recovery and treatment response not significantly different. Exercise group had significantly fewer RBC transfusions and fewer attempts at stem cell collection | Not applicable | Not applicable | (29) |
| Randomized | 24 | Adults (ages 42-74) | Aerobic and Strength | Feasibility of home exercise program (injuries, adherence) | No injuries in either exercise or control group | Lean Body Weight, muscle strength, fatigue, mood disturbances, sleep time | Increased lean body weight in exercise group (+0.40kg /month vs. - 0.44kg /month in control) | (30) |
| Randomized | 42 | Adults (ages 18-65) | Aerobic and Strength | Severity of 5 symptoms clusters: gastrointestinal, cognitive, affective, functional, mucositis | Intervention group mean symptom severity scores were significantly lower in all categories except for affective | Symptom burden | Confirmed that patients undergoing allo-HCT experience a considerable symptom burden | (31) |
| Randomized | 131 | Adult (ages 18-75) | Aerobic and Strength | Physical performance and self-reported physical function | Statistically significant increase in knee extension, walking speed, and 6MWT were found in the exercise group. Grip strength was non-significant | Body composition, Quantified walking activity, self-reported physical activity, fatigue and QOL | No statistical significance | (32) |
| Multicenter randomized | 105 | Adult (ages 18-71) | Aerobic and strength | Fatigue | Up to 15% improvement in exercise group vs up to 28% worsening in control | Physical functioning, anxiety, global distress, 6MWT | Exercise group was found to have significantly higher physical functioning, improved anxiety and 6MWT. Decreased global distress. | (33) |
| Randomized | 67 | Adult (ages 19-73) | Strength | Muscle strength, fatigue, functional ability | Fatigue: significant improvement in intervention group<br>Functional ability: significant increase in timed stair climb & timed up and go<br>Muscle strength: no significant increase, but trend toward improving muscle strength in intervention group | Cross-section area of R. femoris, objective physical activity, quality of life | Cross-section of R. femoris: no significance, in quality of life: no significant difference<br>Physical activity: significant increase in objective physical activity in intervention group | (34) |
| Multicenter randomized | 109 | Adults (ages 19-67) | Aerobic and strength | Physical Fitness: VO2peak, Wpeak, grip strength, 30s chair stand test | No significant benefits in intervention group on physical fitness and fatigue | BMI, sum of 4 skinfolds, max isometric torque of M. quadriceps, HRQOL, symptoms scales of EORTC Myeloma module, distress via HADS questionnaire, subjective and objective physical activity | No significant effects found in secondary outcome measures | (35) |
| Randomized | 20 | Pediatric (ages 4-16) | Aerobic and strength | Immune cell recovery | No difference between groups | Body composition (weight, BMI, Body fat, FFM) | Significant effect across all anthropometric measures (i.e. intervention group increased BMI) | (36) |
| Randomized | 12 | Adults (ages 16-64) | Aerobic and strength | QOL (CARES questionnaire) | Improved QOL in exercise group when compared to pre-transplant and non-exercise group | Peak aerobic capacity | Peak aerobic capacity and QOL were correlated ($p < .05$) | (37) |
| Prospective, repeated measures design | 10 | Adults (ages 48-71) | Strength | Feasibility of exercise based on week to week exercise frequency of participates | Over 6 week period, number of patients exercising 3+ times a week went from 2/10 to 6/10 | Strength (hand grip strength, time needed to stand up, and 30 second chair to stand test). General Health Perceptions via EORTC QLQ-C30 | Strength: hand grip strength (32.66±12.34 to 28.18 ±12.42); time needed to stand 4.63±1.65 to 4.05±1.5; 7.6±3.1 to 8.22±3.42; QOL: 23.51±3.65 to 24±3.88 | (38) |
| Prospective feasibility study | 16 | Adults (ages 60-70) | Aerobic and Strength | More than 50% of patients complete more than 50% of scheduled exercise days during hospitalization | only 15% of scheduled sessions missed by patients | Strength: isometric knee extensor strength, trunk strength; Endurance: 6MWT; QOL; Fatigue; Depression | Equal lower extremity strength, negative trend in endurance capacity, mixed regard in trunk strength | (39) |

In conclusion, frailty is a clinically recognized but underdiagnosed syndrome after allogeneic HCT. A personalized strength training intervention, conducted along with a caregiver in a community setting, can help address some of the root causes of frailty, increasing lean muscle mass and improving self-efficacy. Larger studies are needed to refine our community-based strength training program for allogeneic HCT survivors and their caregivers prior to widespread implementation.

## Data Availability

Data will be posted to ClinicalTrials.gov once the study is complete.

## Acknowledgement of Research Support

This study was funded by the University Of Minnesota Department Of Medicine Women’s Early Research Career program (SGH) and Marrow on the Move, University of Minnesota (SGH).

## REFERENCES

1. M. Arora et al., Physiologic Frailty in Nonelderly Hematopoietic Cell Transplantation Patients: Results From the Bone Marrow Transplant Survivor Study. JAMA Oncol 2, 1277–1286 (2016).

2. L. P. Fried et al., Frailty in older adults: evidence for a phenotype. J Gerontol A Biol Sci Med Sci 56, M146–156 (2001).

3. M. D. Hladek et al., High coping self-efficacy associated with lower odds of prefrailty/frailty in older adults with chronic disease. Aging Ment Health, 1–7 (2019).

4. M. P. Engelen, B. S. van der Meij, N. E. Deutz, Protein anabolic resistance in cancer: does it really exist? Curr Opin Clin Nutr Metab Care 19, 39–47 (2016).

5. E. F. Binder et al., Effects of exercise training on frailty in community-dwelling older adults: results of a randomized, controlled trial. J Am Geriatr Soc 50, 1921–1928 (2002).

6. C. B. Ferreira et al., Effects of a 12-Week Exercise Training Program on Physical Function in Institutionalized Frail Elderly. J Aging Res 2018, 7218102 (2018).

7. D. Langer et al., Exercise training after lung transplantation improves participation in daily activity: a randomized controlled trial. Am J Transplant 12, 1584–1592 (2012).

8. K. R. Lorig, D. S. Sobel, P. L. Ritter, D. Laurent, M. Hobbs, Effect of a self-management program on patients with chronic disease. Eff Clin Pract 4, 256–262 (2001).

9. S. Morishita et al., Prevalence of sarcopenia and relevance of body composition, physiological function, fatigue, and health-related quality of life in patients before allogeneic hematopoietic stem cell transplantation. Support Care Cancer 20, 3161–3168 (2012).

10. M. Pamukcuoglu et al., Hematopoietic Cell Transplant-Related Toxicities and Mortality in Frail Recipients. Biol Blood Marrow Transplant, (2019).

11. T. A. Gooley et al., Reduced mortality after allogeneic hematopoietic-cell transplantation. N Engl J Med 363, 2091–2101 (2010).

12. S. Hashmi, P. Carpenter, N. Khera, A. Tichelli, B. N. Savani, Lost in transition: the essential need for long-term follow-up clinic for blood and marrow transplantation survivors. Biol Blood Marrow Transplant 21, 225–232 (2015).

13. E. H. Wagner, Chronic disease management: what will it take to improve care for chronic illness? Eff Clin Pract 1, 2–4 (1998).

14. T. M. Damush, A. Perkins, K. Miller, The implementation of an oncologist referred, exercise self-management program for older breast cancer survivors. Psychooncology 15, 884–890 (2006).

15. S. Lawn et al., Is self-management feasible and acceptable for addressing nutrition and physical activity needs of cancer survivors? Health Expect 18, 3358–3373 (2015).

16. R. McCorkle et al., Self-management: Enabling and empowering patients living with cancer as a chronic illness. CA Cancer J Clin 61, 50–62 (2011).

17. B. C. Risendal et al., Meeting the challenge of cancer survivorship in public health: results from the evaluation of the chronic disease self-management program for cancer survivors. Front Public Health 2, 214 (2014).

18. S. I. Mishra et al., Exercise interventions on health-related quality of life for people with cancer during active treatment. Clin Otolaryngol 37, 390–392 (2012).

19. S. D. Neupert, M. E. Lachman, S. B. Whitbourne, Exercise self-efficacy and control beliefs: effects on exercise behavior after an exercise intervention for older adults. J Aging Phys Act 17, 1–16 (2009).

20. A. Bandura, Health promotion by social cognitive means. Health Educ Behav 31, 143–164 (2004).

21. S. S. Daniali, F. M. Darani, A. A. Eslami, M. Mazaheri, Relationship between Self-efficacy and Physical Activity, Medication Adherence in Chronic Disease Patients. Adv Biomed Res 6, 63 (2017).

22. A. J. Applebaum et al., A scoping review of caregiver burden during allogeneic HSCT: lessons learned and future directions. Bone Marrow Transplant 51, 1416–1422 (2016).

23. E. D. Hacker et al., Strength training following hematopoietic stem cell transplantation. Cancer Nurs 34, 238–249 (2011).

24. B. A. Cunningham et al., Effects of resistive exercise on skeletal muscle in marrow transplant recipients receiving total parenteral nutrition. JPEN J Parenter Enteral Nutr 10, 558–563 (1986).

25. S. Wallek et al., Impact of the initial fitness level on the effects of a structured exercise therapy during pediatric stem cell transplantation. Pediatr Blood Cancer 65, (2018).

26. M. Jarden, M. T. Baadsgaard, D. J. Hovgaard, E. Boesen, L. Adamsen, A randomized trial on the effect of a multimodal intervention on physical capacity, functional performance and quality of life in adult patients undergoing allogeneic SCT. Bone Marrow Transplant 43, 725–737 (2009).

27. E. A. Coleman et al., Effects of exercise on fatigue, sleep, and performance: a randomized trial. Oncol Nurs Forum 39, 468–477 (2012).

28. M. L. Shelton et al., A randomized control trial of a supervised versus a self-directed exercise program for allogeneic stem cell transplant patients. Psychooncology 18, 353–359 (2009).

29. E. A. Coleman et al., Effects of exercise in combination with epoetin alfa during high-dose chemotherapy and autologous peripheral blood stem cell transplantation for multiple myeloma. Oncol Nurs Forum 35, E53–61 (2008).

30. E. A. Coleman et al., Feasibility of exercise during treatment for multiple myeloma. Cancer Nurs 26, 410–419 (2003).

31. M. Jarden, K. Nelausen, D. Hovgaard, E. Boesen, L. Adamsen, The effect of a multimodal intervention on treatment-related symptoms in patients undergoing hematopoietic stem cell transplantation: a randomized controlled trial. J Pain Symptom Manage 38, 174–190 (2009).

32. R. H. Knols et al., Effects of an outpatient physical exercise program on hematopoietic stem-cell transplantation recipients: a randomized clinical trial. Bone Marrow Transplant 46, 1245–1255 (2011).

33. J. Wiskemann et al., Effects of a partly self-administered exercise program before, during, and after allogeneic stem cell transplantation. Blood 117, 2604–2613 (2011).

34. E. D. Hacker et al., Strength Training to Enhance Early Recovery after Hematopoietic Stem Cell Transplantation. Biol Blood Marrow Transplant 23, 659–669 (2017).

35. S. Persoon et al., Randomized controlled trial on the effects of a supervised high intensity exercise program in patients with a hematologic malignancy treated with autologous stem cell transplantation: Results from the EXIST study. PLoS One 12, e0181313 (2017).

36. C. Chamorro-Vina et al., Exercise during hematopoietic stem cell transplant hospitalization in children. Med Sci Sports Exerc 42, 1045–1053 (2010).

37. S. Hayes, P. S. Davies, T. Parker, J. Bashford, B. Newman, Quality of life changes following peripheral blood stem cell transplantation and participation in a mixed-type, moderate-intensity, exercise program. Bone Marrow Transplant 33, 553–558 (2004).

38. E. D. Hacker, J. L. Larson, D. Peace, Exercise in patients receiving hematopoietic stem cell transplantation: lessons learned and results from a feasibility study. Oncol Nurs Forum 38, 216–223 (2011).

39. M. K. Schuler et al., Feasibility of an exercise programme in elderly patients undergoing allogeneic stem cell transplantation - a pilot study. Eur J Cancer Care (Engl) 25, 839–848 (2016).

